# Factors Influencing Low-Acuity Emergency Medical Services Use: An Observational Study Guided by the Andersen Behavioral Model

**DOI:** 10.64898/2026.02.19.26346664

**Authors:** Hannah C Muthersbaugh, James E Winslow, Joseph M Grover, Chris M Gillette

## Abstract

**Objectives:** Emergency Medical Services (EMS) demand is increasing, with a growing proportion of low-acuity encounters. Prior studies are limited by regional sampling, inconsistent definitions, narrow observation periods, and limited theoretical grounding. The objective of this study was to identify predisposing, enabling, and need-based factors associated with EMS transport among low-acuity emergency department (ED) patients, guided by Andersen’s Behavioral Model of Health Services Use.

**Methods:** We conducted a secondary, retrospective observational study using a 10% random sample of multicenter electronic health record (EHR) data from 21 emergency departments in the southeastern United States. To be eligible to be included in the analysis, the visit had to be for: (1) patient age >17 years of age, (2) occur between January 1, 2016, to April 29, 2025, (3) triaged as Emergency Severity Index (ESI) 4 or 5, and (4) ended in a final visit disposition of being discharged. The primary outcome was EMS utilization. Independent variables were categorized as predisposing, enabling, or need-based factors according to Andersen’s model. We used multivariable logistic regression to estimate adjusted odds ratios (aORs) with 95% confidence intervals.

**Results:** Among 41,772 low-acuity ED encounters, 3,233 (7.7%) arrived by EMS. Increased odds of EMS use were associated with older age (per 10-year increase; aOR 1.30, 95% CI=1.27-1.33), male sex (aOR 1.20, 95% CI=1.12-1.30), being retired or disabled (aOR 3.60, 95% CI=3.15-4.10), being unemployed (aOR 2.26, 95% CI=2.04-2.52), having a nighttime presentation (aOR 1.63, 95% CI=1.51-1.76), and mental health diagnosis (aOR 1.76, 95% CI=1.62-1.90). Protective factors included White race (aOR=0.89, 95% CI=0.83-0.96), established primary care (aOR=0.57, 95% CI=0.57-0.62), weekend presentation (aOR 0.91; CI = 0.84-0.99), and visits during (aOR 0.63, 95% CI=0.55-0.71) or after (aOR 0.54, 95% CI=0.48-0.61) the COVID-19 period. Rurality, insurance, and primary language were not associated with EMS use.

**Conclusions:** Predisposing and enabling factors were the predominant drivers of low-acuity EMS utilization in this sample. Expanding access to primary care and behavioral health services, especially for older patients, may reduce EMS use for low-acuity complaints while preserving EMS capacity for higher-acuity emergencies.

## Introduction

From its formal inception in 1966 under the Department of Transportation, Emergency Medical Services (EMS) in the United States were designed with a specific mission: rapid patient transport to hospital care (1, 2). This transport-centric structure, rooted in the Anglo-American “scoop-and-run” model (3), is reinforced by federal reimbursement systems that link payments to hospital transport rather than to treatment (4). While effective for time-sensitive emergencies, routine use of the scoop-and-run model for low-acuity complaints may contribute to ongoing emergency department (ED) overcrowding, prolonged wall times, and decreased EMS capacity for high-acuity complaints (5–7). At the same time, recent national reports indicate that the EMS workforce faces substantial strain, with rising call volumes, high staffing turnover, and shortages of EMS clinicians that hinder timely emergency response (8). These system pressures have been shown to exacerbate delays in EMS and ED care and diminish readiness for critical cases, particularly as low-acuity transports consume limited resources that are already being stretched thin (5, 6, 9). National estimates suggest that up to 15% of Medicare-covered EMS transports are likely non-emergent and can be treated in outpatient settings (4).

EMS agencies nationwide are confronting escalating call volumes, workforce shortages, and a growing burden of low-acuity encounters (5, 6, 10, 11). Recent research suggests that sociodemographic and access-related factors, including social vulnerability, transportation barriers, older age, unemployment or disability, limited availability of primary care, and perceived lack of alternative care options, influence low-acuity EMS utilization (12–15). Significant gaps remain in the literature, including reliance on single-EMS-agency or municipality-level analyses, inconsistent definitions of “low-acuity,” and short observation periods, often one to three years, with COVID-era studies typically focused on discrete pandemic intervals rather than longitudinal patterns (12, 16–19).

Andersen’s Behavioral Model (ABM) posits that health service utilization is shaped by three domains: predisposing factors, enabling factors, and need-based factors (20, 21). The model has been widely applied using administrative claims data in a variety of health care settings and consistently demonstrates that utilization is driven not only by clinical need but also by the interaction of individual characteristics, resource availability, and social context (22, 23). Despite this, few studies explicitly apply this model to EMS research, limiting understanding of how structural and social determinants collectively shape ambulance use for low-acuity complaints and potentially obscuring modifiable system-level drivers. To address this critical gap in the literature, this study aims to examine predisposing, enabling, and need-based factors associated with EMS transport among low-acuity ED patients across ED systems in the southeastern US.

## Methods

We conducted a secondary, retrospective observational study using multicenter electronic health record (EHR) data from 21 emergency departments that deliver care across a multi-hospital regional referral network serving more than 7 million people in North Carolina, South Carolina, Georgia, and Virginia (24). The Wake Forest University School of Medicine Institutional Review Board approved this study with a waiver of informed consent for retrospective analysis of de-identified data (IRB00122446). We used the Strengthening the Reporting of Observational Studies in Epidemiology (STROBE) statement to guide study reporting (*Appendix A*). We used de-identified data and integrated clinical information from legacy and current EHR systems. An experienced data analyst extracted records from June to September 2025, capturing visits from January 1, 2016, to April 29, 2025, across multiple EHR platforms.

### Participants

At all EDs, triage nurses used the validated five-level Emergency Severity Index (ESI), in which ESI 4 and 5 are categorized as non-urgent (12, 25). The two datasets initially included all emergency department visits triaged only as ESI 4 or 5 between January 1, 2016, and April 29, 2025. The dataset contained 878,220 records representing 543,763 unique patients. ESI 4 and 5 encounters accounted for 21.6% of the total number of patients evaluated during this period. We used a 10% random sample to reduce computational burden and the likelihood of selection bias affecting the study’s findings.

To be included in the analysis, visits had to be: (1) from January 1, 2016, to April 29, 2025, and (2) triaged as ESI 4 or 5. We excluded ED visits involving patients who were (a) less than 18 years of age, (b) currently pregnant, or (c) presenting with suicidal ideation, involuntarily committed (IVC), or self-injurious behavior. We also excluded nonstandard modes of arrival (e.g., air medical services, sheriff, and rapid response).

We defined low-acuity presentation as triage ESI 4 or 5 and discharge from the emergency department (25). Previous estimates indicate that approximately 5.5% of patients triaged as low-acuity eventually require acute hospitalization (26); therefore, we excluded patients triaged as ESI 4 or 5 who were subsequently admitted for low-acuity presentations.

### Measures

We defined the primary outcome as arrival at the ED via EMS (1 = arrived by EMS, 0 = arrived by another mode, e.g., car/private vehicle or taxi).

We applied Andersen’s Behavioral Model of Health Services Utilization (ABM) to guide the selection of independent variables (**Figure 1)**. The ABM posits that a complex mix of predisposing, enabling, and need factors influences patient use of health services (23, 27). The ABM characterizes predisposing factors as underlying patient characteristics that affect the propensity to use health services, such as demographic and social variables. In this study, we defined predisposing factors as age, sex, race, primary language, and employment status. Enabling factors are the logistical and structural resources that facilitate or impede access to care, including insurance type, rurality, having an established primary care clinician, and time of arrival (weekend or nighttime presentation). Need-based factors capture both perceived and evaluated illness severity and clinical need for care; for this analysis, these included mental health comorbidities and the COVID-19 Pandemic. Together, these domains provide a structured, theory-driven approach for identifying the determinants of EMS utilization among low-acuity patients.

**Figure 1:**
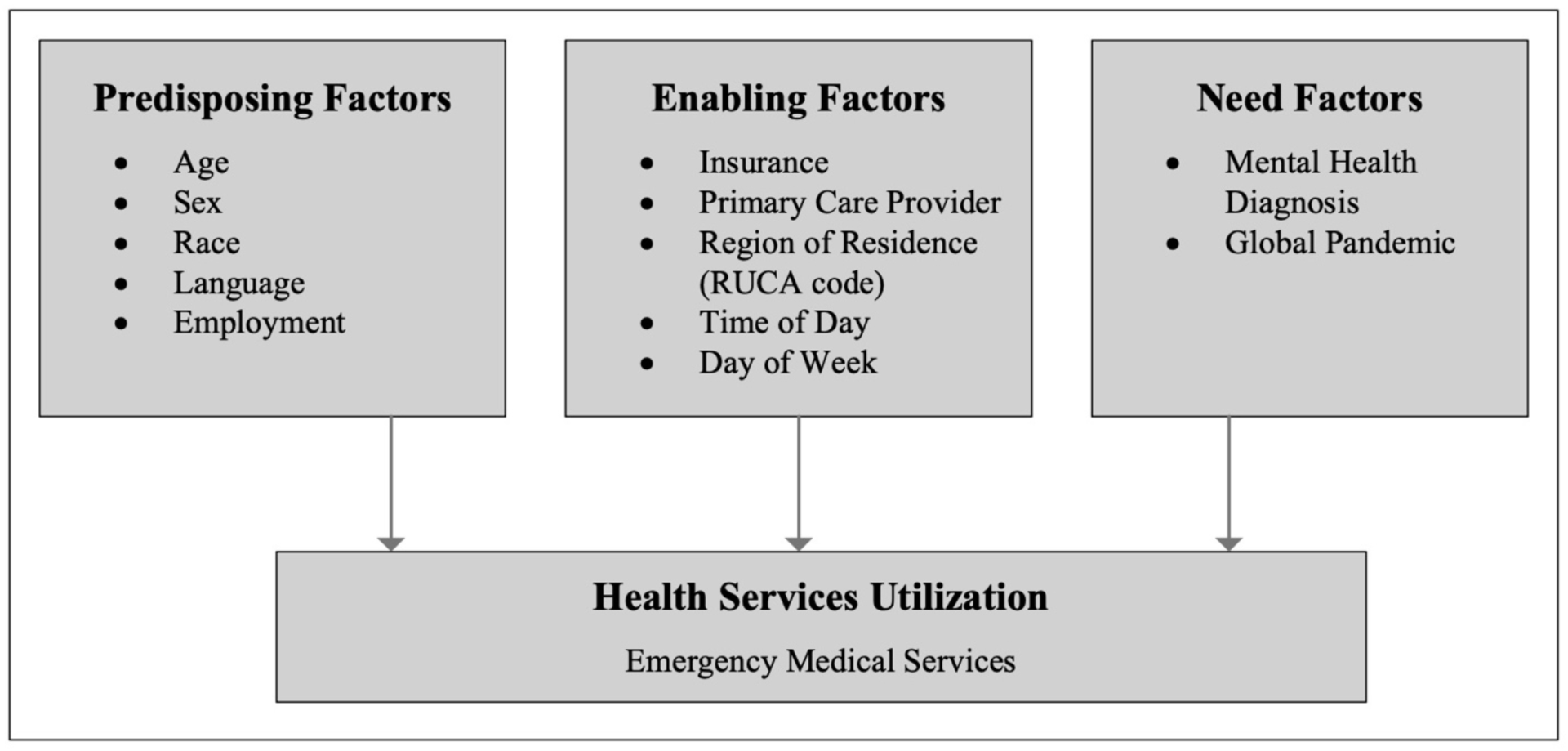
Andersen’s Behavioral Model of Health Services Utilization.

We defined the following variables as dichotomous: low acuity presentation of ESI 4 or 5 with ED discharge (1 = Yes; 0 = No), mode of arrival (1 = ambulance; 0 = all other modes [car, public transportation, taxi, walk-in, and wheelchair]), visit day (1 = weekend [Saturday/Sunday]; 0 = weekday [Monday-Friday]), sex (1 = Male; 0 = Female), primary language (1 = English; 0 = non-English), established primary care clinician (1 = yes; 0 = no or unknown), presence of a mental health diagnosis identified by ICD-10 “F” codes (1 = yes; 0 = no), and time of arrival (1 = day [7 am-6:59 pm]; 0 = [7 pm-6:59 am]). Rurality was defined using Rural-Urban Commuting Area (RUCA) codes and dichotomized according to the WWAMI Rural Health Research Center guidelines (1 = urban [RUCA 1.0-3.0]; 0 = rural [RUCA 4.0-10.6]) (28). Race was initially evaluated as a categorical variable but was recoded into a dichotomous variable (1 = White, 0 = Non-White) because the Non-White category had small subgroup sizes, presumably reflecting heterogeneity in how race is coded during ED visits, a limitation noted in prior studies demonstrating variability between recorded and self-identified race and ethnicity in clinical data (29).

We analyzed financial class, employment status, and pandemic period as categorical variables. Financial class was categorized as (1 = Commercial/Private/Third-party [Blue Cross Blue Shield, managed care, liability]; 2 = Medicaid/Medicare/Government [Medicaid, Medicare Advantage, CHAMPVA, TRICARE, and other government programs]; 3 = Self-pay/None [self-pay, financial counseling]). Employment status was categorized as (1 = Employed [including active military]; 2 = Not working with regular income [disabled, retired]; 3 = Unemployed/Unknown). The pandemic period was categorized as (1 = Pre-COVID [January 1, 2016–March 10, 2020]; 2 = COVID [March 11, 2020–May 11, 2023]; 3 = Post-COVID [May 12, 2023–April 29, 2025]).

Age was analyzed as a continuous variable, scaled in 10-year increments to facilitate interpretation.

To mitigate inconsistent data capture and information bias, we harmonized variable definitions across the two EHRs. For example, one EHR recorded ambulance as the mode of arrival, whereas the new EHR recorded each ambulance by its county of affiliation. We applied similar harmonizations to arrival by car and to Medicaid variants.

### Statistical Analysis

We managed data using SAS v9.4 (Cary, NC) and conducted analyses in SPSS v30.0.0.0.0 (Chicago, IL). We used listwise deletion to handle missing data. We summarized continuous variables using means and standard deviations when the data were normally distributed, and medians and interquartile ranges when the data were non-normally distributed. We presented categorical variables as frequencies and percentages. To assess bivariate associations among the independent variables in the ABM, we used chi-squared tests, ANOVA, or independent samples t-tests, as appropriate. We identified independent variables with significant pairwise associations (p < 0.05) and considered them as potential confounders. We evaluated multicollinearity using variance inflation factors (VIFs), with values >5 indicating collinearity that could bias the multivariable regression model (30). All VIF values were < 5, indicating low multicollinearity and supporting the model’s stability. We then constructed a multivariable logistic regression model with EMS use for low-acuity visits (1 = ambulance; 0 = other modes of arrival) as the dependent variable. We selected independent variables *a priori* using Andersen’s Behavioral Model, including predisposing factors (age, sex, race, primary language), enabling factors (insurance coverage, employment status, primary care clinician, day of week), and need factors (mental health diagnosis, COVID era). To improve the interpretability of age as a predictor, we rescaled age to 10-year increments for the regression. For all analyses, we used an ɑ<0.05 to indicate statistical significance.

## Results

**Figure 2** illustrates cohort selection. The final analytic sample included 41,772 low-acuity emergency department encounters. Among these, 3,233 (7.7%, n=3,233) arrived by EMS, and 38,539 (92.3%, n=38,539) arrived by other modes (private vehicle, walk-in, or public transport).

**Figure 2:**
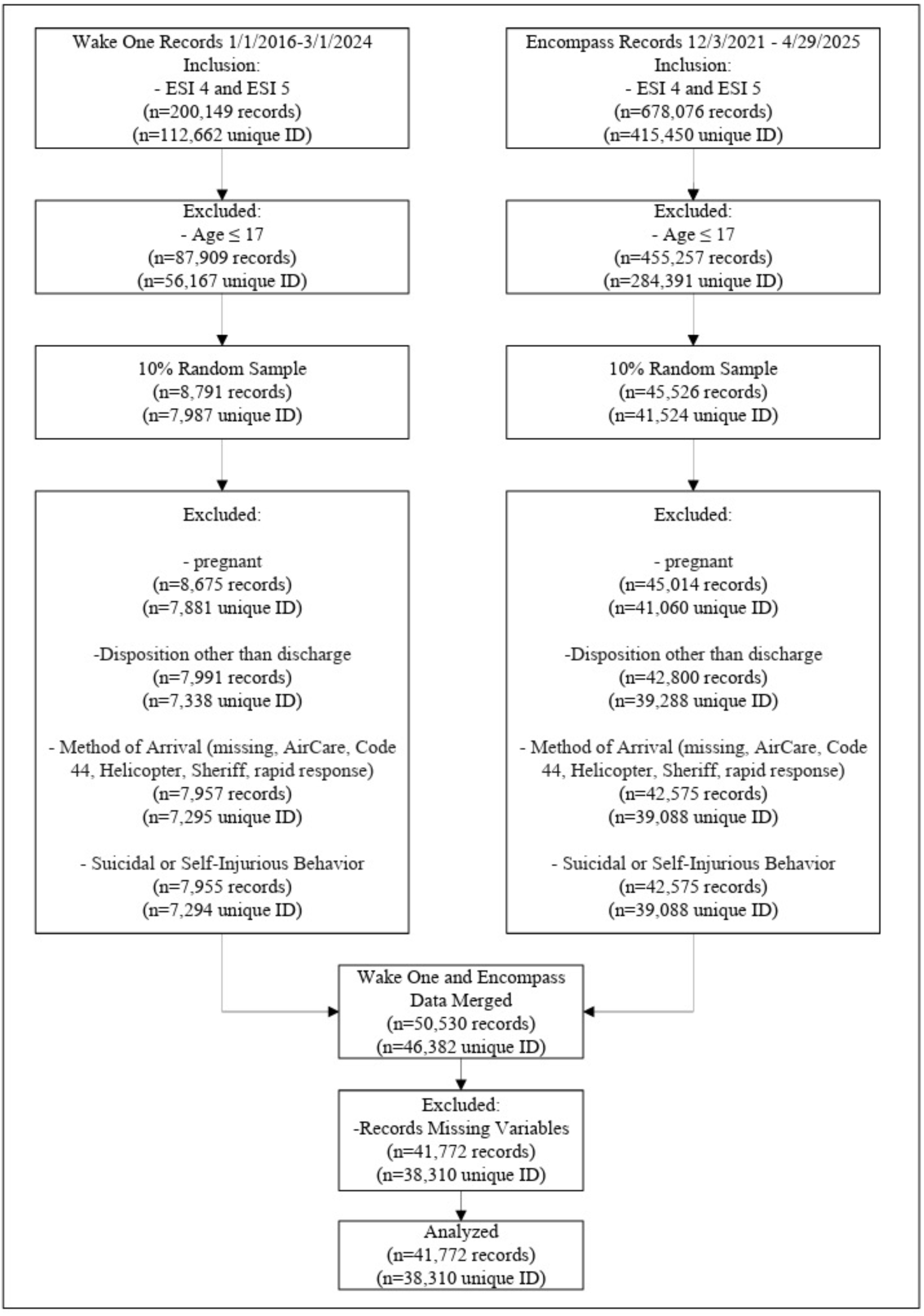
Cohort Selection.

**Table 1** summarizes baseline demographic, predisposing, enabling, and need characteristics. Patients transported by EMS were generally older, more likely to be male, unemployed, or retired/disabled, and less likely to have an established primary care clinician than those who did not use EMS for low-acuity events.

**Table 1:** Sociodemographic Characteristics of Sample (n=41,772)

| Independent Variable | Used EMS<br>(n=3233) | Did not use EMS<br>(n=38539) | P Value |
| --- | --- | --- | --- |
| **Predisposing** |  |  |  |
| Age | **52.0 (21.29)** | **42.7 (18.17)** | **<0.001** |
| Sex |  |  |  |
| Male | **48.7 (1576)** | **42.9 (16535)** | **<0.001** |
| Female | **51.3 (1657)** | **57.1 (22040)** |  |
| Race |  |  |  |
| White | 48.6 (1572) | 47.2 (18186) | 0.116 |
| Non-White | 51.4 (1661) | 52.8 (20353) |  |
| Language |  |  |  |
| English | 97.0 (3135) | 96.7 (37258) | 0.371 |
| Non-English | 3.0 (98) | 3.3 (1281) |  |
| Employment |  |  |  |
| Employed | **22.1 (716)** | **50.5 (19470)** | **<0.001** |
| Not Working, Regular Income | **47.8 (1546)** | **19.9 (7660)** |  |
| Unemployed | **30.0 (971)** | **29.6 (11403)** |  |
| **Enabling** |  |  |  |
| Insurance |  |  |  |
| Commercial/Private/3rd Party | **28.2 (913)** | **45.1 (17372)** | **<0.001** |
| Medicare/Medicaid/Government | **71.7 (2318)** | **54.8 (21111)** |  |
| Self-Pay/None | **0.1 (2)** | **0.1 (56)** |  |
| Primary Care Clinician |  |  |  |
| Yes | **67.5 (2181)** | **70.9 (27328)** | **<0.001** |
| No | **32.5 (1052)** | **29.1 (11211)** |  |
| Region of Residence |  |  |  |
| Urban | 83.1 (2686) | 82.5 (31795) | 0.424 |
| Rural | 16.9 (547) | 17.5 (6740) |  |
| Time of Day |  |  |  |
| 7a-7p | **57.2 (1850)** | **67.2 (25893)** | **<0.001** |
| 7p-7a | **42.8 (1383)** | **32.8 (12646)** |  |
| Day of Week |  |  |  |
| Weekend | 27.3 (883) | 28.7 (11062) | 0.093 |
| Weekday | 72.7 (2350) | 71.3 (27477) |  |
| **Need** |  |  |  |
| Mental Health Diagnosis |  |  |  |
| No | **51.7 (1673)** | **69.0 (26592)** | **<0.001** |
| Yes | **48.3 (1560)** | **31.0 (11947)** |  |
| COVID |  |  |  |
| Pre-COVID | **13.4 (434)** | **8.0 (3083)** | **<0.001** |
| COVID | **29.0 (939)** | **28.8 (11117)** |  |
| Post-COVID | **57.5 (1860)** | **63.2 (24339)** |  |

We performed a multivariable logistic regression with EMS usage (vs. other modes of arrival) as the dependent variable. Independent variables included age, sex, race, primary language, insurance, employment, day of week, time of day, RUCA code, established primary care clinician, mental health diagnosis, and COVID era. **Table 2** presents adjusted odds ratios (aORs) for the multivariable model. **Supplemental Table 1** presents unadjusted odds ratios.

**Table 2:** Multivariable Logistic Regression Predicting Low-Acuity EMS Use (n=41,772)

| **Independent Variable** | **aOR\*** | **95% CI** | **p-value** |
| --- | --- | --- | --- |
| Age (age/10) | **1.30** | **1.27–1.33** | **<0.001** |
| Sex (Male) | **1.20** | **1.12–1.30** | **<0.001** |
| Race (White) | **0.89** | **0.83–0.96** | **0.004** |
| Language (English) | 0.95 | 0.77–1.18 | 0.647 |
| Employment (Retired/Disabled vs Employed) | **3.60** | **3.15–4.10** | **<0.001** |
| Employment (Unemployed vs Employed) | **2.26** | **2.04–2.52** | **<0.001** |
| Insurance (Government vs Commercial) | 1.03 | 0.93–1.13 | 0.594 |
| Insurance (Self-pay vs Commercial) | 0.45 | 0.11–1.90 | 0.279 |
| Primary Care Clinician (Yes) | **0.57** | **0.57–0.62** | **<0.001** |
| RUCA (Rural) | 0.99 | 0.90–1.10 | 0.853 |
| Time of Day (7p–7a) | **1.63** | **1.51–1.76** | **<0.001** |
| Visit Day (Weekend) | **0.91** | **0.84–0.99** | **0.021** |
| Mental Health Diagnosis (Yes) | **1.76** | **1.62–1.90** | **<0.001** |
| COVID (COVID) | **0.63** | **0.55–0.71** | **<0.001** |
| COVID (Post-COVID) | **0.54** | **0.48–0.61** | **<0.001** |

\*aORs adjusted for age, sex, race, primary language, employment status, insurance type,

Older patients (aOR=1.30, 95% CI=1.27-1.33), those who are male (aOR=1.20, 95% CI=1.12-1.30), being retired/disabled (aOR=3.60, 95% CI=3.15-4.10), or unemployed (aOR=2.26, 95% CI=2.04-2.52), going to the ED between 7p-7a (aOR=1.63, 95% CI=1.51-1.76), and having a mental health diagnosis (aOR=1.76, 95% CI=1.62-1.90) significantly increased the likelihood of using EMS for transport to the ED.

Being of White race (aOR=0.89, 95% CI=0.83-0.96), visiting the ED on the weekend (aOR=0.91, 95% CI=0.84-0.99), having an established primary care clinician (aOR=0.57, 95% CI=0.57-0.62), and visiting the ED during COVID (aOR=0.63, 95% CI=0.55-0.71) and post-COVID (aOR=0.54, 95% CI=0.48-0.61) were significantly associated with decreased likelihood of using EMS for transport to the ED. There was no statistically significant association (p>0.05) between EMS use and primary language, rural area, or insurance type.

## Discussion

To our knowledge, this is the first multi-state study examining EMS utilization for low acuity patient ED encounters. We found that low-acuity EMS utilization is shaped by a complex mix of predisposing, enabling, and need-related factors, consistent with Andersen’s Behavioral Model. In older adults, EMS utilization may reflect underlying functional limitations that increase reliance on EMS transport, rather than greater perceived urgency (31). Prior work also shows increasing rates of transport for falls and medical complaints among patients aged 65 years and older (32). While increasing age was associated with a cumulative rise in low-acuity EMS use in this cohort, unemployment and retirement/disability were also independently associated with substantially higher odds of low-acuity EMS utilization, indicating that social vulnerability may exert a stronger influence than aging alone (14, 15).

Having an established primary care clinician and presenting to the ED on weekends were associated with reduced odds of EMS utilization, as were visits during or after the COVID-19 pandemic. Our findings align with previous studies that have identified a lack of timely access to primary care as a driver of EMS utilization (17). Prior studies also report lower overall EMS and ED utilization on weekends, followed by a sharp increase early in the week, suggesting that factors beyond outpatient access influence EMS use (33). Low-acuity transports declined during and after the COVID-19 pandemic, consistent with national trends (18, 34). Public health messaging, concerns about infection, and the rapid expansion of telehealth likely contributed to reduced EMS utilization during this period (34–38). At the same time, reduced in-person primary care visits may have shifted some patients towards EMS use while remaining protective at the population level (19, 39).

White race and presenting to the ED on weekends were associated with a modest protective effect; however, the limited magnitude suggests they are unlikely to be primary drivers of EMS utilization. Primary language, rurality, and insurance type were not significantly associated with EMS use for low-acuity ED visits.

These findings align with prior studies that identify age, unemployment, disability, and limited access to primary care as key drivers of low-acuity EMS use in the United States and internationally (12–15). The prevalence in our cohort (7.7%) aligns with national estimates, indicating that 5-15% of ambulance transports in the United States may be non-emergent or potentially avoidable (4, 9, 13). This represents a substantial share of EMS utilization, with significant implications for system capacity (40).

Our findings, consistent with prior work, highlight the need for multi-modal interventions rather than reliance on a single approach. Community Paramedic models and nurse triage phone lines have demonstrated the ability to divert demand away from traditional EMS and ED pathways, with some Community Paramedic models reducing utilization by approximately 50% (41, 42). Insurance status was not associated with EMS use for low-acuity ED visits, suggesting a potential funding source for alternative means of reaching these patients, should reimbursement models permit it. Expanding access to primary care, including APP-led models, may further address lower-acuity needs outside the emergency setting and lessen reliance on EMS (43). While much of the research explores treatment-in-place strategies to reduce transports (44–46), recent work linking dispatch categories to time-critical interventions and improved telephone triage supports the development of programs that identify and redirect low-acuity patients earlier in the care pathway (10, 42).

As EMS quality frameworks expand beyond response intervals to include appropriate resource use and transport destination decisions (40), policy and reimbursement models must address these broader drivers of care-seeking behavior. Innovations that decouple payment from hospital transport, such as the CMS Emergency Triage, Treat, and Transport (CMS ET3) Model (47), exemplify approaches that align resource allocation with patient needs (4, 11, 48). Persistent challenges such as limited primary care access, mental health instability, and socioeconomic vulnerability underscore the need for integrated public health, behavioral health, and primary care strategies to reduce low-acuity EMS use (49).

## Limitations

Our study has several limitations to generalizability. The retrospective, observational design precludes causal inference, and reliance on data from a single system and limited EHR types may limit generalizability to regions with different EMS structures. This concern is partially mitigated by the health system’s broad geographic reach across multiple states and diverse practice settings. Although a 10% random sample reduced computational burden, the lack of stratification by site or year may have introduced selection bias; however, fully random selection makes meaningful changes in effect size unlikely. Extracted EHR data are subject to misclassification, including triage level, transport mode, insurance status, primary care establishment, and race, which are documented through a combination of patient self-report and staff entry in the electronic record. Race was analyzed dichotomously (White vs. Non-White) because small subgroup sizes precluded stable categorical comparisons, thereby limiting the ability to examine heterogeneity among racial and ethnic groups and potentially obscuring differences within these populations (**Supplemental Table 2**). Variability in ESI assignment may also contribute to measurement error. Mental health diagnoses were identified using ICD-10 code “F” codes, which do not distinguish condition acuity or severity. We excluded missing data rather than imputing values, potentially biasing results toward complete records. Finally, unmeasured factors such as insecurity regarding food, transportation, and housing, prior EMS use, access to unscheduled outpatient care, and other comorbidities were not captured and may also influence EMS utilization. Patient complexity, including ambulation status, was also not captured in the dataset for all patients. Future research should investigate how other need variables, such as comorbidities, influence EMS utilization.

## Conclusions

In conclusion, predisposing and enabling factors were the predominant drivers of low-acuity EMS utilization in this cohort. Older age, male sex, unemployment, retirement or disability, nighttime presentation, and the presence of a mental health diagnosis were associated with increased odds of EMS transport, whereas established primary care, weekend presentation, and visits during or after the COVID-19 period were associated with lower use. Geographic residence, insurance type, and primary language were not associated with EMS use. Within the limits of the variables examined, these findings suggest that social and behavioral determinants most strongly influence low-acuity EMS utilization within this health system.

These results support the continued development of community interventions and reimbursement reforms that better align EMS response with patient needs while preserving system readiness for higher-acuity emergencies. Future work should evaluate strategies that expand primary care access, strengthen mental health and community paramedicine resources, integrate telehealth-supported navigation, and align reimbursement models with treatment-in-place and alternative destination pathways to reduce low-acuity EMS use and improve care continuity. In a landscape in which EMS agencies are increasingly expected to balance efficiency, equity, and preparedness, targeting the underlying social drivers of EMS use will be critical to achieving sustainable, patient-centered system performance.

## Data Availability

Data are not publicly available due to institutional data use agreements, but may be available from the corresponding author upon reasonable request and institutional approval. Data sharing policies for the Wake Forest School of Medicine are available here: https://ctsi.wakehealth.edu/service/scientific-editing-services.

## Acknowledgements

This manuscript was created as part of graduation requirements for the DMSc program of Wake Forest University School of Medicine Physician Assistant Studies. The authors would like to acknowledge Dr. Kelly R. Conner, PhD, PA-C, FAES, and Dr. Natalie Smith, DMSc, MS, PA-C, for their assistance with editorial review and manuscript preparation. Their contributions improved the manuscript’s clarity and organization but did not meet the criteria for authorship.

## Declaration of Generative AI in Scientific Writing

The authors did not use a generative artificial intelligence (AI) tool or service to assist with the preparation or editing of this work. The author(s) take full responsibility for the content of this publication.

## Authorship Statement

HM conceived and designed the study, conducted the data analysis, interpreted the results, and drafted the manuscript, serving as the primary author and lead contributor across all aspects of the work. CG contributed to study design, assisted with data management and analytic strategy, participated in interpretation of findings, and critically revised the manuscript for important intellectual content. JW and JG critically reviewed the manuscript for important intellectual context and provided subject-matter expertise regarding interpretation and application within the field of emergency medical services. All authors approved the final version of the manuscript and agree to be accountable for all aspects of the work.

